# A rare haplotype of the *GJD3* gene segregating in familial Meniere Disease interferes with connexin assembly

**DOI:** 10.1101/2024.01.16.24300842

**Authors:** Alba Escalera-Balsera, Paula Robles-Bolivar, Alberto M. Parra-Perez, Silvia Murillo-Cuesta, Han Chow Chua, Lourdes Rodríguez-de la Rosa, Julio Contreras, Ewa Domarecka, Juan Carlos Amor-Dorado, Andrés Soto-Varela, Isabel Varela-Nieto, Agnieszka J Szczepek, Alvaro Gallego-Martinez, Jose A. Lopez-Escamez

**Affiliations:** Otology & Neurotology Group CTS495, Instituto de Investigación Biosanitaria, ibs.GRANADA, Universidad de Granada, 18071 Granada, Spain; Division of Otolaryngology, Department of Surgery, Universidad de Granada, Granada, Spain; Sensorineural Pathology Programme, Centro de Investigación Biomédica en Red en Enfermedades Raras, CIBERER, Madrid, Spain; Institute for Biomedical Research Sols-Morreale (IIBm), Spanish National Research Council-Autonomous University of Madrid (CSIC-UAM), Madrid, Spain; Rare Diseases Networking Biomedical Research Centre on Rare Diseases (CIBERER), Carlos III Institute of Health, Madrid, Spain; La Paz Hospital Institute for Health Research (IdiPAZ), Madrid, Spain; Sydney Pharmacy School, Faculty of Medicine and Health and Charles Perkins Centre, The University of Sydney, Sydney, New South Wales, Australia; Anatomy and Embryology Department, Faculty of Veterinary, Universidad Complutense de Madrid, Madrid, Spain; Department of Otorhinolaryngology, Head and Neck Surgery, Charité-Universitätsmedizin Berlin, Corporate Member of Freie Universität Berlin and Humboldt-Universität zu Berlin, Charitéplatz 1, 10117 Berlin, Germany; Department of Otolaryngology, Hospital Can Misses, Ibiza, Spain; Division of Otoneurology, Department of Otorhinolaryngology, Complexo Hospitalario Universitario, Santiago de Compostela, Spain; Department of Surgery and Medical-Surgical Specialities, Universidade de Santiago de Compostela, Santiago de Compostela, Spain; Meniere’s Disease Neuroscience Research Program, Faculty of Medicine & Health, School of Medical Sciences, The Kolling Institute, University of Sydney, Sydney, New South Wales, Australia

**Keywords:** Meniere Disease, Hearing loss, Inner ear, Tectorial membrane, Exome sequencing, Bioinformatics, Connexin gene, Connexin protein, Protein modeling, Immunohistofluorescence

## Abstract

Familial Meniere Disease (FMD) is a rare polygenic disorder of the inner ear. Mutations in the connexin gene family, which encodes gap junction proteins, can also cause hearing loss, but their role in FMD is largely unknown. Here, we found an enrichment of rare missense variants in the *GJD3* gene when comparing allelic frequencies in FMD (N=94) with the Spanish reference population (OR=3.9[1.92-7.91], FDR=2.36E-03). In the *GJD3* sequence, we identified a rare haplotype (TGAGT) composed of two missense, two synonymous, and one downstream variants. This haplotype was found in five individuals with FMD, segregating in three unrelated families with a total of ten individuals; and in another eight Meniere Disease individuals. *GJD3* encodes the gap junction protein delta 3, also known as human connexin 31.9 (CX31.9). The protein model predicted that the NP_689343.3:p.(His175Tyr) missense variant could modify the interaction between connexins and the connexon assembly, affecting the homotypic GJD3 gap junction between cells. Our studies in mice revealed that the mouse ortholog *Gjd3* - encoding Gjd3 or mouse connexin 30.2 (Cx30.2) - was expressed in the organ of Corti and vestibular organs, particularly in the tectorial membrane, the base of inner and outer hair cells and the nerve fibers. The present results describe a novel association between *GJD3* and familial FMD, providing evidence that FMD is related to changes in the inner ear channels; in addition, it supports a new role of tectorial membrane proteins in FMD.

## 1. Introduction

Connexins are essential plasma membrane proteins in epithelial intercellular junctions ^1^. Connexin protein subunits form a hexameric complex named connexon, and each connexon forms a hemichannel in the plasma membrane. The arrangement of two connexons between adjacent cells forms a gap junction, which communicates the cytoplasm of both cells and controls the intercellular exchange of small molecules, metabolites and ions ^2,3^. In the inner ear, gap junctions are essential in maintaining the fluid’s homeostasis. They are located in the organ of Corti and the lateral wall of the cochlea, including the stria vascularis ^2,4,5^.

The tectorial membrane (TM) is an extracellular matrix located along the length of the organ of Corti, where the lateral surface is attached to the stereocilia of the mechanosensory hair cells. It intercedes in the deflection of the stereocilia, being involved in the hair cell stimulation and, therefore, in the gating of channels ^6,7^.

Meniere Disease (MD, MIM: 156000) is an inner ear disorder characterized by episodic vertigo and associated with sensorineural hearing loss (SNHL), tinnitus and/or aural fullness ^8^. The criteria to diagnose MD are based on the clinical symptoms occurring during the attacks of vertigo and the documentation of SNHL by pure tone audiogram before, during, or after the episode of vertigo. Several subgroups of patients with MD have been reported according to associated co-morbidities ^9^, such as migraine or autoimmune disorders, cytokine profile ^10^ or methylation signature ^11^.

The syndrome shows familial aggregation and several rare variants in different genes have been reported in singular families, manifesting a considerable genetic heterogeneity ^12^. Furthermore, exome sequencing in additional families with MD supports a burden of rare variation in three central SNHL genes in familial MD (FMD), including *OTOG* (MIM: 604487) ^13^, *MYO7A* (MIM: 276903) ^14^ and *TECTA* (MIM: 602574) ^15^.

Furthermore, genetic studies have demonstrated the importance of some connexins expressed in the inner ear for human hearing. *GJB2* (MIM: 121011) mutations lead to approximately half of monogenic non-syndromic SNHL, besides *GJB6* (MIM: 604418) and *GJB3* (MIM: 603324) mutations cause non-syndromic HL ^2,3^. In this way, Gallego-Martinez et al. ^16^ found a significant overload of missense variants in *GJB2* in sporadic MD (SMD) patients that were not found in the reference population.

We have sequenced and analyzed a large cohort of patients with MD and identified a rare haplotype TGAGT in the gene *GJD3* (MIM: 607425), segregating the phenotype in multiple families, supporting *GJD3* as a new gene associated with FMD.

## 2. Materials and methods

### 2.1. Patient selection

Patients were diagnosed as definite MD according to the diagnostic criteria described by the International Classification Committee for Vestibular Disorders of the Barany Society ^17^. A total of 94 FMD patients from Spanish referral centers belonging to 70 different families with one or more first-degree relatives affected by MD were included. In addition, a dataset of 313 patients with SMD was studied. Pure-tone audiograms were retrieved to assess hearing loss (HL), and audiometry was represented using tidyr ^18^, ggplot2 ^19^, dplyr ^20^, ggpubr ^21^, scales ^22^ R packages.

The human ethics protocol of this study was approved by the Institutional Review Board (Protocol number: 1805-N-20), and all the subjects signed a written informed consent to donate biological samples. The animal experiments were approved by the Governmental Ethics Commission for Animal Welfare in Berlin, Germany (LaGeSo Berlin, Germany; approval number: T 0235/18) and by the Dirección General de Agricultura, Ganadería y Alimentación in Comunidad de Madrid, Spain (approval number PROEX 325.4/21). The investigation followed the principles of the Declaration of Helsinki revised in 2013 ^23^.

### 2.2. Exome sequencing

To perform WES, blood samples were obtained from each patient. DNA samples were extracted using prepIT-L2P (DNA Genotek, Ottawa, Canada) and QIAamp DNA Mini Kit (Qiagen, Venlo, The Netherlands) following manufacturer’s protocols. The quality controls required for exome sequencing were performed as previously described ^24^. DNA libraries were prepared to select coding regions by using SureSelect Human All Exon V6 capture kit (Agilent Technologies, Santa Clara, CA, USA) and paired-end sequenced on the Illumina HiSeq 4000 platform with a mean coverage of 100X.

### 2.3. Dataset generation

Paired-end sequences were mapped to the GRCh38/hg38 human reference genome, using the maximal exact matches algorithm Burrows-Wheeler Aligner. Nextflow Sarek v2.7.1 workflow, included in nf-core ^25^, was utilized to perform the exome reference alignment, base quality score recalibration (BSQR), variant calling, and quality filtering. Duplicated reads were removed, and the alignment quality was evaluated ^26^. Genetic variants were called using the Haplotypecaller function, from GATK ^27^. In this stage, Single Nucleotide Variants (SNVs) and short insertions and deletions (indels) were detected at nucleotide resolution, and the results were saved in a Variant Calling Format (VCF) file for each subject.

The VCF files were normalized with the norm function from bcftools ^28^. Each VCF file was filtered according to the criteria followed to create the gnomAD database: Allele balance (AB) ≥ 0.2 and AB ≤ 0.8 (for heterozygous genotypes only), genotype quality (GQ) ≥ 20, and depth (DP) ≥ 10 (5 for haploid genotypes on sex chromosomes) ^29^. Using the merge function of bcftools, a MD variant dataset containing the variants of all the individuals was generated ^28^. Following GATK best practices, a variant quality filtering was carried out with Variant Score Recalibration (VQSR), which calculates a new quality score: VQSLOD. Variants that accomplished a VQSLOD < 90 were retained.

### 2.4. Variant annotation and prioritization strategy

Variants included in the dataset were annotated using Ensembl Variant Effect Predictor (VEP). Then, variants in connexin genes for SMD and FMD were selected and saved separately for further analyses^3,30^.

Two independent databases were used to retrieve the allelic frequencies (AF) of the variants in three reference populations. The AF for Non-Finish European (NFE, n = 32,299) and global population (n = 71,702) were obtained from the gnomAD database v.3.0 ^23^. Population-specific AF for the Spanish population were retrieved from the Collaborative Spanish Variant Server (CSVS, n = 2,048) ^32^. For this, we performed a liftover from GRCh19/hg19 to GRCh38/hg38 reference genomes, which only included SNVs.

To perform the Gene Burden Analysis (GBA), an AF < 0.05 was selected as a threshold in the three databases. Besides, variants were classified according to the consequence in the protein to perform 6 different GBA (missense; frameshift, inframe deletion and inframe insertion; stop gain; 3’UTR; 5’UTR; and synonymous) for familial patients (Figure S1). Only one individual from each family was selected, whenever possible, according to the lowest age of onset and/or from the last generation.

To search genes associated with FMD, a GBA was carried out in familial cases. The aggregated AF for each gene calculated for the three reference populations (gnomAD NFE, gnomAD global and CSVS) was compared with the aggregated AF in FMD, and odds ratios (OR) with 95% confidence interval (CI) were calculated. Furthermore, two-sided p-values were obtained and corrected according to the False Discovery Rate (FDR) for multiple testing by the total number of variants identified for each gene; and Etiological Fraction (EF) was calculated, as previously described ^33^. Genes with an adjusted *p*-value < 0.05 and OR ≥ 1 in one of the three comparisons with each reference population were considered enriched.

To prioritize those genes obtained as enriched in the GBA, the dataset RNA-Seq in P0 from the murine cochlea to contrast HC with the rest of the cochlear duct from the gene Expression Analysis Resource (gEAR) database ^34^ was used. Genes expressed in the inner ear were selected for further analysis. Variants in selected genes were assessed by the Combined Annotation Dependent Depletion (CADD) ^35^ score and following the standards and guidelines described by the American College of Medical Genetics and Genomics (ACMG) and the Association for Molecular Pathology (AMP) ^36^.

Visual inspection confirmed candidate variants in BAM files to rule out false positives. Moreover, the variants NC_000017.11:g.40363293G>A, NC_000017.11:g.40363579G>T and NC_000017.11:g.40363294C>G in *GJD3* were validated by Sanger sequencing, using the following primers: CCACCGCGAAATAGAAGAGC (Fw) and AGGACGAGCAAGAGGAGTTC (Rv).

The constraint metrics were obtained from the gnomAD database v.2.1.1 ^31^. The ratio of the observed/expected missense variants and the Z score were calculated with the deviation of observed from the expected were considered in this study. A Z score calculated by the ratio between observed variation and expected depletion of variation at a 1kb scale.

### 2.5. Linkage disequilibrium and haplotype

The complete list of *GJD3* variants was downloaded from gnomAD database v.3.1.2 ^31^ to calculate the linkage disequilibrium (LD) among all known variants in *GJD3*. The R^2^ score was obtained and represented for each pair of variants, using the *LDmatrix* and *LDheatmap* function from the LDlinkR ^37^ and LDheatmap ^38^ R packages, respectively. Besides, the *LDhap* function from the LDlinkR R package was used to calculate the haplotype frequencies of shared variants in the global (ALL), European (EUR) and Iberian in Spain (IBS) populations. Population genotype data used in LDlinkR was obtained from Phase 3 (Version 5) of the 1,000 Genomes Project.

### 2.6. Computational protein modeling

The GJD3 amino acid sequence was retrieved from Uniprot (Q8N144). The monomer structural model was predicted using AlphaFold2 ^39^. The structural model of the homomeric connexon, with a C6 symmetry, and the homotypic gap junction (two connexons) was predicted using HDOCK ^40^. HDOCK does not use the entire protein in the docking process, focusing on predicting the conformation of the binding sites of the protein to reduce the computational cost. The quality of the protein structural models was assessed using the structure validation algorithms Molprobity ^41^, Verify3D ^42^, ERRAT ^43^, ProSA-web ^44^, and QMEANDisCo ^45^. The mutated GJD3 protein was modeled by comparative homology modeling with MODELLER 10.4 ^46^ using the wild-type GJD3 protein model as a template. These in-silico models were used to predict the protein stability change (ΔΔG) caused by the candidate variants, using the ENCoM ^47^, DynaMut2 ^48^, I-Mutant ^49^, mCSM ^50^, mCSM-membrane ^51^, mmCSM-PPI ^52^, SDM^53^ and, PremPS ^54^ tools. Variants were classified as neutral when −0.5 < ΔΔG_pred_ < 0.5 ^55^.

The GJD3 structural model was submitted to the ModelArchive database (https://modelarchive.org/doi/10.5452/ma-bwdwf; Public access after publication. Temporary access code: Uy6tl2t305).

### 2.7. Mouse cochlear RNA Isolation and quantitative RT-PCR

Cochlear samples from C57BL/6JCrl mice of 1, 6 and 12 months of age were obtained and processed as reported ^56,57^. Immediately after dissection, cochleae were frozen in RNAlater® solution (Ambion, Foster City, CA, USA). Cochlear RNA was extracted using the RNeasy Plus Mini kit (Qiagen, Hilden, Germany) automated on the Qiacube (Qiagen, Hilden, Germany). RNA integrity was assessed with an Agilent 2100 bioanalyzer (Agilent Technologies, Santa Clara, CA, USA). cDNA was generated from pooled cochlear RNA extracts (each pool included three cochleae from different animals per age group) by reverse transcription with the High-Capacity cDNA Reverse Transcription Kit (Applied Biosynthesis, Thermo Fisher Scientific) and amplified in triplicate by real time quantitative PCR (RT-qPCR) in a QuantStudio 7 Flex PCR System (Applied Biosystems, Foster City, CA, USA) using gene-specific primers (Fw: CGCACACGGTCGACTGTTT, Rv: GCGAAGTAGAAGACCACGAAGAC). Data was collected after each amplification step and analyzed with QuantStudio™ Real-Time PCR software 1.3 (Applied Biosystems). *Hprt1* (hypoxanthine phosphoribosyltransferase 1) and *Tbp* (TATA-Box Binding Protein) genes were used as housekeeping genes for normalization. Differences between ages were calculated by the Student’s t-test with ΔCt values (Ct target gene - Ct value reference gene), and the Fold Change (FC) and Standard error of the mean (SEM) were obtained following the 2^−ΔΔCT^ method. Outliers were identified by the interquartile range (IQR) method filtering those values 1.5 IQR below the first quartile or 1.5 IQR above the third quartile. The statistics and visualizations were performed using ggplot2 ^19^ and ggpubr ^21^ R packages.

### 2.8. Mouse cochlear Immunohistofluorescence

Cochlear samples from postnatal day 3 (P3), postnatal day 30 (P30), and postnatal day 90 (P90) C57BL/6JCrl mice were obtained and processed following standard protocols, as reported ^56,57^. Briefly, adult mice were perfused with 4% paraformaldehyde in PBS. Dissected inner ears were post-fixed with 4% paraformaldehyde, decalcified in 5% EDTA, cryopreserved with sucrose and embedded in a medium for cryotomy. For immunostaining, cryostat cross sections (10 μm) were firstly dried at room temperature and then washed with PBS 0.1 M. After that, specimens were incubated for 1 hour in a normal goat serum solution in a humidified chamber at room temperature to block nonspecific binding sites. Then the specimens were incubated for 24 hours with primary antibody (rabbit polyclonal anti-Connexin 30.2, LifeTechnologies, # 40-7400) diluted (1:125) in goat serum/PBS/Triton X-100 at 4°C in a humidified chamber. On the following day, after 4x 15 minutes of washing, specimens were incubated with the specific fluorophore-conjugated antibody (Alexa Fluor 488 Goat Anti-Rabbit IgG (H+L), LifeTechnologies, # A11034) diluted (1:400) in goat serum/PBS/Triton X-100 for 1.5 hours at room temperature in a humidified chamber. Finally, the specimens were coverslipped using ProLong® Gold Antifade Reagent with DAPI (Cell Signaling Technologies, Danvers, MA, USA, # 8961). The fluorescent images of stained cryosections from P90 mice were taken with an epifluorescence microscope (Nikon 90i, Tokyo, Japan); and from P3 and P30 mice with a Leica SPE confocal microscope. The confocal images were merged in a z-stack using ImageJ ^58^.

### 2.9. Functional validation in Xenopus oocytes

Human CX31.9 wild-type (WT), NP_689343.3:p.(His175Tyr) and NP_689343.3:p.(Arg253Pro) complementary DNAs (cDNAs) cloned between NheI and BamHI sites in a modified pCDNA3.1(+) vector containing 5′ and 3′-*Xenopus* globin UTR and a polyadenylation signal, were generated using custom gene synthesis with codon optimization for *Homo sapiens* (GenScript). For expression in *Xenopus laevis* oocytes, plasmid DNAs were linearized with BamHI restriction enzyme, from which capped RNAs were synthesized using the T7 mMessage mMachine Kit (Invitrogen).

Oocyte extraction from *Xenopus laevis* frogs was performed following a protocol approved by the Animal Ethics Committee of The University of Sydney (AEC No. 2016/970) in accordance with the National Health and Medical Research Council of Australia code for the care and use of animals. Ovarian lobes were sliced into small pieces using surgical knives and defolliculated by collagenase treatment. Healthy-looking stage V-VI oocytes were injected with 50 nL of a 0.5 ng/nL RNA and incubated at 18 °C in modified Barth’s solution (96 mM NaCl, 2.0 mM KCl, 1 mM MgCl_2_, 1.8 mM CaCl_2_, 5 mM HEPES, 2.5 mM sodium pyruvate, 0.5 mM theophylline, and 100 μg/mL gentamicin; pH 7.4). Two to three days after RNA injection, two-electrode voltage-clamp measurements were performed on oocytes continuously perfused in recording solution mimicking the endolymph (100 mM KCl, 2 NaCl, 1.8 mM BaCl_2_, 5 mM HEPES; pH 7.4) at room temperature using an Axon GeneClamp 500B amplifier (Molecular Devices, LLC, Sunnyvale, CA, USA). Data were acquired using the pCLAMP 10 software (Molecular Devices) and a Digidata 1440A digitizer (Molecular Devices), sampled at 10 kHz. Recording microelectrodes with resistances around 0.2–1.0 MΩ were pulled from borosilicate glass capillaries (Harvard Apparatus) using a PC-100 Dual-Stage Glass Micropipette Puller (Narishige) and were filled with 3 M KCl.

## 3. Results

### 3.1. Overload of missense variants in the *GJD3* gene in familial cases

A total of 71 variants in 19 connexin genes with an AF < 0.05 were retained to carry out a GBA in FMD individuals (Table 1, Figure S1, Table S1). Variants were classified according to the consequence in the protein, the most common being missense variants. We found an enrichment of missense variants in the *GJD3* gene when comparing the AF in FMD against the Spanish population from CSVS (OR = 3.9 [1.92-7.91], FDR = 2.36E-03, EF = 0.74). Moreover, an enrichment of synonymous variants in the *GJD3* gene comparing the AF in the FMD against the Spanish population from CSVS (OR = 3.46 [1.89-6.33], FDR = 5.76E-04, EF = 0.71), and also the global population from gnomAD (OR = 2.9 [1.63-5.14], FDR = 3.02E-03, EF = 0.65).

**Table 1.** Connexins genes with enrichment of rare variants (Allelic Frequency (AF) < 0.05) in Familial Meniere Disease (FMD).

| Consequence(s) | Gene symbol | Protein | Number variants | Number individuals | gnomAD NFE |  |  | gnomAD |  |  | CSVS |  |  |
| --- | --- | --- | --- | --- | --- | --- | --- | --- | --- | --- | --- | --- | --- |
|  |  |  |  |  | OR [CI] | FDR | EF | OR [CI] | FDR | EF | OR [CI] | FDR | EF |
| Missense | *GJA1* | CX43 | 1 | 5 | 2.36 [0.96-5.78] | 8.99E-01 | 0.58 | 4.15 [1.7-10.15] | 2.75E-02 | 0.76 | 2.12 [0.84-5.34] | > 1 | 0.53 |
|  | *GJA3* | CX46 | 3 | 4 | 4.43 [1.65-11.91] | 4.79E-02 | 0.77 | 5.83 [2.17-15.65] | 7.00E-03 | 0.83 | 6.71 [2.26-19.91] | 8.48E-03 | 0.85 |
|  | *GJA5* | CX40 | 1 | 1 | 28.12 [3.72-212.83] | 1.85E-02 | 0.96 | 39.3 [5.3-291.32] | 4.92E-03 | 0.97 | 2.51 [0.32-19.42] | > 1 | 0.60 |
|  | *GJA10* | CX62 | 1 | 1 | 159.42 [16.48-1542.37] | 1.78E-04 | 0.99 | 265.31 [29.46-2389.29] | 9.69E-06 | 1.00 | - | - | - |
|  | *GJB5* | CX31.1 | 3 | 3 | 102.47 [29.34-357.95] | 6.04E-12 | 0.99 | 3.6 [1.15-11.22] | 4.11E-01 | 0.72 | 12.65 [3.26-49.08] | 3.43E-03 | 0.92 |
|  | *GJB7* | CX25 | 1 | 1 | 15.42 [2.09-113.78] | 1.09E-01 | 0.94 | 20.4 [2.8-148.64] | 4.38E-02 | 0.95 | 5.86 [0.68-50.52] | > 1 | 0.83 |
|  | *GJD3* | CX31.9 | 2 | 5 | 1.45 [0.75-2.82] | > 1 | 0.31 | 2.19 [1.13-4.26] | 3.10E-01 | 0.54 | 3.9 [1.92-7.91] | 2.36E-03 | 0.74 |
| Frameshift, inframe deletion and insertion | *GJD3* | CX31.9 | 1 | 1 | 473.13 [29.44-7603.58] | 4.14E-05 | 1.00 | 69.73 [9.15-531.62] | 1.26E-04 | 0.99 | - | - | - |
| 3'UTR | *GJC3* | CX30.2 | 1 | 1 | - | - | - | - | - | - | 29.46 [1.83-473.51] | 1.70E-02 | 0.97 |
| Synonymous | *GJA4* | CX37 | 1 | 1 | - | - | - | 1061.55 [66.06-17059.72] | 9.63E-06 | 1.00 | - | - | - |
|  | *GJB3* | CX31 | 2 | 1 | 13.66 [3.33-55.97] | 2.81E-03 | 0.93 | 0.29 [0.07-1.18] | 9.32E-01 | -2.40 | 3.35 [0.77-14.49] | > 1 | 0.70 |
|  | *GJC2* | CX47 | 1 | 8 | 1.43 [0.7-2.92] | > 1 | 0.30 | 2.42 [1.19-4.96] | 1.67E-01 | 0.59 | 4.04 [1.89-8.61] | 3.02E-03 | 0.75 |
|  | *GJD3* | CX31.9 | 4 | 7 | 1.92 [1.08-3.41] | 2.58E-01 | 0.48 | 2.9 [1.63-5.14] | 3.02E-03 | 0.65 | 3.46 [1.89-6.33] | 5.76E-04 | 0.71 |
|  | *GJE1* | CX23 | 2 | 2 | 238.73 [43.54-1308.91] | 2.85E-09 | 1.00 | 32.57 [7.93-133.69] | 1.47E-05 | 0.97 | 10.12 [1.95-52.4] | 5.80E-02 | 0.90 |

The AF in each reference database was compared to the AF in the FMD cohort: the Non-Finnish European for gnomAD (gnomAD NFE), the global population for gnomAD (gnomAD), and the Spanish population for CSVS (CSVS). All the samples are heterozygous for the variants. OR: Odd ratio; CI: Confidence Interval; FDR: p-value corrected by False Discovery Ratio; EF: Etiological Fraction.

### 3.2. Segregation of a rare haplotype in the *GJD3* gene with FMD

We found several rare variants in the *GJD3* gene in FMD: two missense variants (NC_000017.11:g.40363058C>G, NC_000017.11:g.40363293G>A) an inframe deletion (NC_000017.11:g.40363099_40363101del), two downstream variants (NC_000017.11:g.40356228C>T, NC_000017.11:g.40356584C>A) and four synonymous variants (NC_000017.11:g.40363294C>G, NC_000017.11:g.40363327G>A, NC_000017.11:g.40363528G>A, NC_000017.11:g.40363579G>T) (Table 2). Interestingly, five of these variants, g.40356228C>T (downstream), g.40363058C>G (missense), g.40363293G>A (missense), g.40363294C>G (synonymous) and g.40363579G>T (synonymous) were shared among five patients with FMD, and were segregated in three families, with a total of ten individuals carrying a haplotype with the five variants. Moreover, this haplotype was found in another eight individuals with non-familial MD that were initially considered as sporadic cases. However, four of them have relatives with incomplete MD phenotype (HL or episodic vestibular symptoms). The variants g.40363293G>A, g.40363294C>G and g.40363579G>T were validated by Sanger sequencing (Figure S2).

**Table 2.** Variants in *GJD3* connexin gene found in Familial Meniere Disease (FMD) cases.

| Variant | Amino acid change | Consequence | CADD score | ACMG classification | Number individuals |  |  | AF |  |  |
| --- | --- | --- | --- | --- | --- | --- | --- | --- | --- | --- |
|  |  |  |  |  | FMD in GBA | FMD total (families segregate) | SMD | gnomAD NFE | gnomAD | CSVS |
| NC_000017.11:g.40356228C>T | - | Downstream | 5.07 | Uncertain significance (PP1,BP4) | 5<sup>a</sup> | 10<sup>a</sup> (3) | 8<sup>a</sup> | 2.30E-02 | 1.53E-02 | 1.40E-02 |
| NC_000017.11:g.40356584C>A | - | Downstream | 7.072 | Likely benign (BP4) | 2 | 2 | 20 | 1.76E-02 | 1.27E-02 | 2.60E-02 |
| NC_000017.11:g.40363058C>G | NP_689343.3:p.(Arg253Pro) | Missense | 7.695 | Likely benign (PP1,BS1,BS2,BP4) | 5<sup>a</sup> | 10<sup>a</sup> (3) | 11<sup>b</sup> | 2.30E-02 | 1.54E-02 | 6.00E-03 |
| NC_000017.11:g.40363099_40363101del | NP_689343.3:p.(Pro248del) | Inframe deletion | 14.95 | Uncertain significance (PM2,BP4) | 1 | 1 | 0 | 1.60E-05 | 1.06E-04 | - |
| NC_000017.11:g.40363293G>A | NP_689343.3:p.(His175Tyr) | Missense | 18.58 | Likely benign (PP1,BS1,BS2,BP4) | 5<sup>a</sup> | 10<sup>a</sup> (3) | 8<sup>a</sup> | 2.30E-02 | 1.53E-02 | 1.10E-02 |
| NC_000017.11:g.40363294C>G | NP_689343.3:p.Pro174= | Synonymous | 16.66 | Likely benign (PP1,BS1,BS2,BP4,BP7) | 5<sup>a</sup> | 10<sup>a</sup> (3) | 8<sup>a</sup> | 2.30E-02 | 1.53E-02 | 1.10E-02 |
| NC_000017.11:g.40363327G>A | NP_689343.3:p.Ala163= | Synonymous | 16.74 | Likely benign (PM2,BP4,BP7) | 1 | 1 | 0 | - | 7.00E-06 | 2.53E-04 |
| NC_000017.11:g.40363528G>A | NP_689343.3:p.Ser96= | Synonymous | 16.77 | Likely benign (BP4,BP7,PM2) | 1 | 1 | 0 | 4.18E-04 | 2.38E-04 | - |
| NC_000017.11:g.40363579G>T | NP_689343.3:p.Leu79= | Synonymous | 20.8 | Likely benign (PP1,BS1,BS2,BP4,BP7) | 5<sup>a</sup> | 10<sup>a</sup> (3) | 8<sup>a</sup> | 2.30E-02 | 1.54E-02 | 1.50E-02 |

<sup>a</sup>: Same individuals with the TGAGT haplotype; <sup>b</sup>: Same 8 individuals with the TGAGT haplotype + another 3 individuals with the CGGCG haplotype. CADD: Combined Annotation Dependent Depletion; ACMG: American College of Medical Genetics and Genomics; FMD: Familial Meniere Disease; GBA: Gene burden analysis; SMD: Sporadic Meniere Disease; gnomAD: Genome Aggregation Database; NFE: Non-Finnish European; CSVS: Collaborative Spanish Variant Server.

The LD study showed a strong correlation between the four shared coding variants. The linkage between g.40363293G>A, g.40363294C>G and g.40363579G>T was complete (R^2^ = 1), and LD between each of them with g.40363058C>G was almost complete (R^2^ = 0.966) (Figure S3).

Since these five variants (g.40356228C>T, g.40363058C>G, g.40363293G>A, g.40363294C>G and g.40363579G>T) seem to form a haplotype (TGAGT), we study the AF in the 1,000 Genomes Project population: 0.0058 for the global population, 0.0159 for the European and 0.0093 for the Iberian in Spain (Table S2). Besides, in the 1,000 Genome Project we found the haplotype CGGCG, present only in one individual in the population of 1,000 Genomes Project, originally from Spain (AF global = 0.0002, AF European = 0.001, AF Iberian in Spain = 0.0047). This CGGCG haplotype had the reference allele for all the variants except for the g.40363058C>G missense variant. This genotype has been shown in another three non-familial MD individuals of our cohort (Table 2, Table S2).

The constraint of the *GJD3* gene for missense variants *-* according to gnomAD - is determined by the Z score = 0.76, and the ratio observed/expect = 0.78 [0.64 - 0.95]. The ratio less than 1 and the positive Z score value suggest that the gene is highly conserved, and intolerant to variation.

### 3.3. Characterization of individuals carrying *GJD3* variants

A detailed clinical characterization of the hearing profile was performed for the 18 individuals with the TGAGT rare haplotype. From the five familial probands, the segregation in three of those families was confirmed (families F1, F2, and F3; pedigrees are not shown to prevent patients’ identification, but they are available for reviewers a researcher upon reasonable request); however, it was not possible to obtain blood samples from relatives of two probands (F4 and F5).

The proband (III-6) of family 1 (F1) was a woman in the early 50s with definite MD. Her progenitor (II-5) was also diagnosed with definite MD with Tumarkin crisis, both carrying the variants studied. Moreover, a relative of the proband (II-1) was diagnosed with probable MD and he did not have the same variants. His clinical history differs from the other two cases, as his age of onset for MD was in the 70s, relatively older than that of the proband and her progenitor, which was in the 30s and in the 60s, respectively. Moreover, his flat hearing profile does not show a drop in high frequencies (Figure 1).

**Figure 1.**
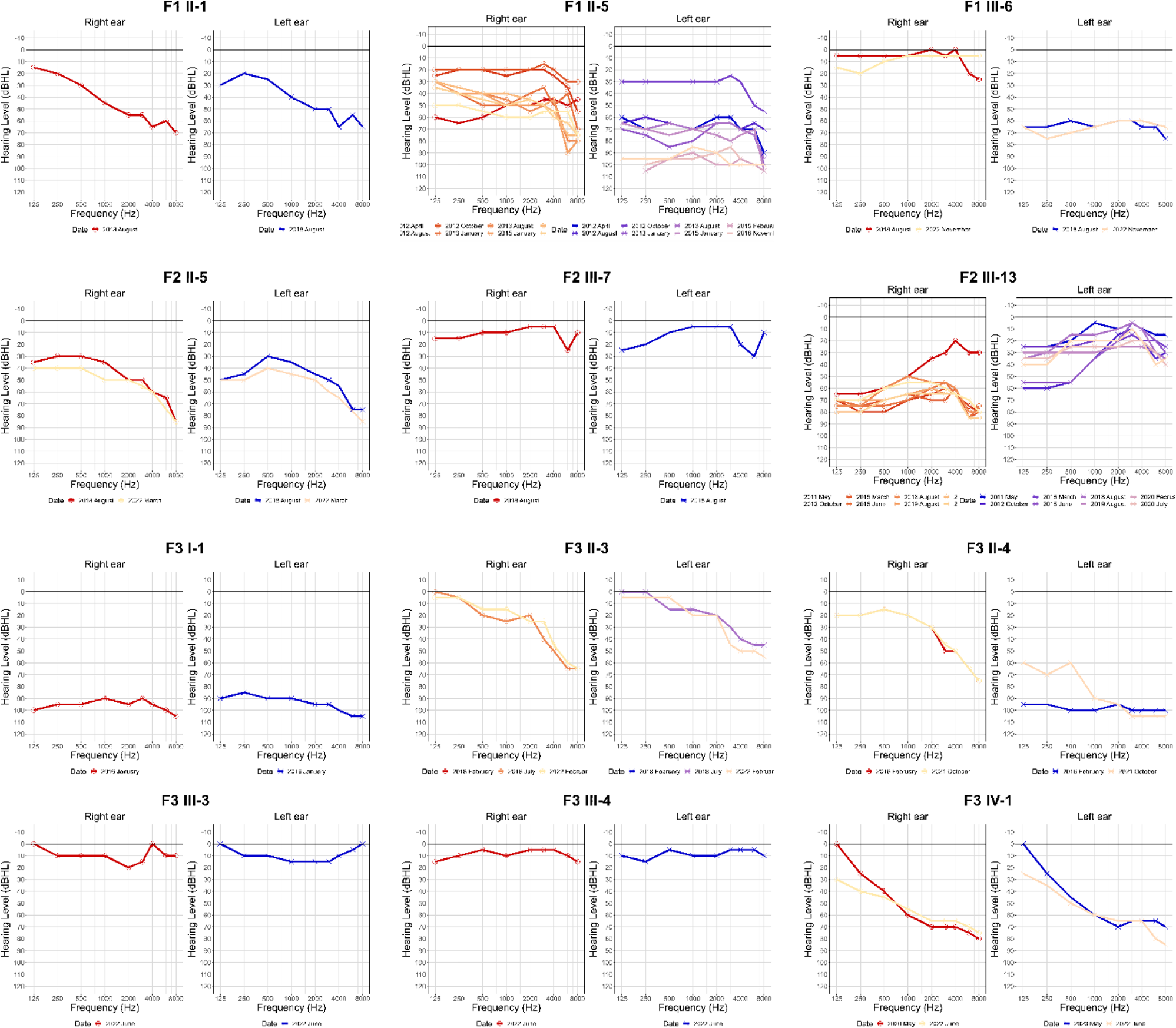
Air conduction audiogram of families 1-3. dB: decibels, kHz: kilohertz.

In family 2 (F2) the proband (III-13) was a woman in the early 60s with definite MD. Her progenitor (II-5) also suffered from MD, and four of her five siblings and two relatives have vertigo attacks. Besides, one of her two relatives presented HL, and his descendant (III-7) was also diagnosed with definite MD. They all started with episodic symptoms at a similar age in their 40s. The three cases with MD have the studied variants. Nevertheless, it was not possible to obtain samples for the individuals with incomplete phenotypes.

The proband (II-4) of the third family (F3) was a man in the early 70s with definite MD. His progenitor (I-1) was also diagnosed with definite MD, whereas his sibling (II-3) had probable MD based on the audiometry results. In this family, all these three individuals carry the variants of interest, and they have a similar age of onset, in their 40s. Furthermore, none of his children suffer from inner ear disorders, but his granddaughter (IV-1) has presented high-frequency HL since her birth. Neither IV-1 nor her parents (III-3 and III-4) were carriers of the studied variants.

The case in F4 was a woman in the 70s with definite MD, bilateral HL (Figure S4), with an onset at her 30s and a history of migraine and autoimmune diseases. The F5 case was a woman with definite MD, with bilateral HL, and her age at HL onset was in her 20s.

In addition, four (S1, S2, S3, and S4) of the eight SMD cases with the studied variants have first-degree relatives with vertigo and SNHL or only episodic vertigo. Interestingly, the three SMD cases (S9, S10 and S11) with the CGGCG haplotype did not report relatives with HL or vestibular disorders (Figure S4).

### 3.4. Protein modeling

The monomeric structure of the Gap junction delta-3 protein (Q8N144) - called GJD3 and CX31.9 - encoded by the *GJD3* gene, was predicted using AlphaFold2. Furthermore, both the hexameric connexon in a closed conformation, with a C6 symmetry, and the structure of the homotypic GJD3 channel, formed by two identical connexons along a two-fold crystallographic symmetry axis, were modeled (Figure 2). Based on the geometrical validation results, reliable models have been built compared to structures solved by experimental methods at the geometrical level (Table S3). These models were used to predict the impact of the variants found on the stability of the monomer, connexon and gap junction models.

**Figure 2.**
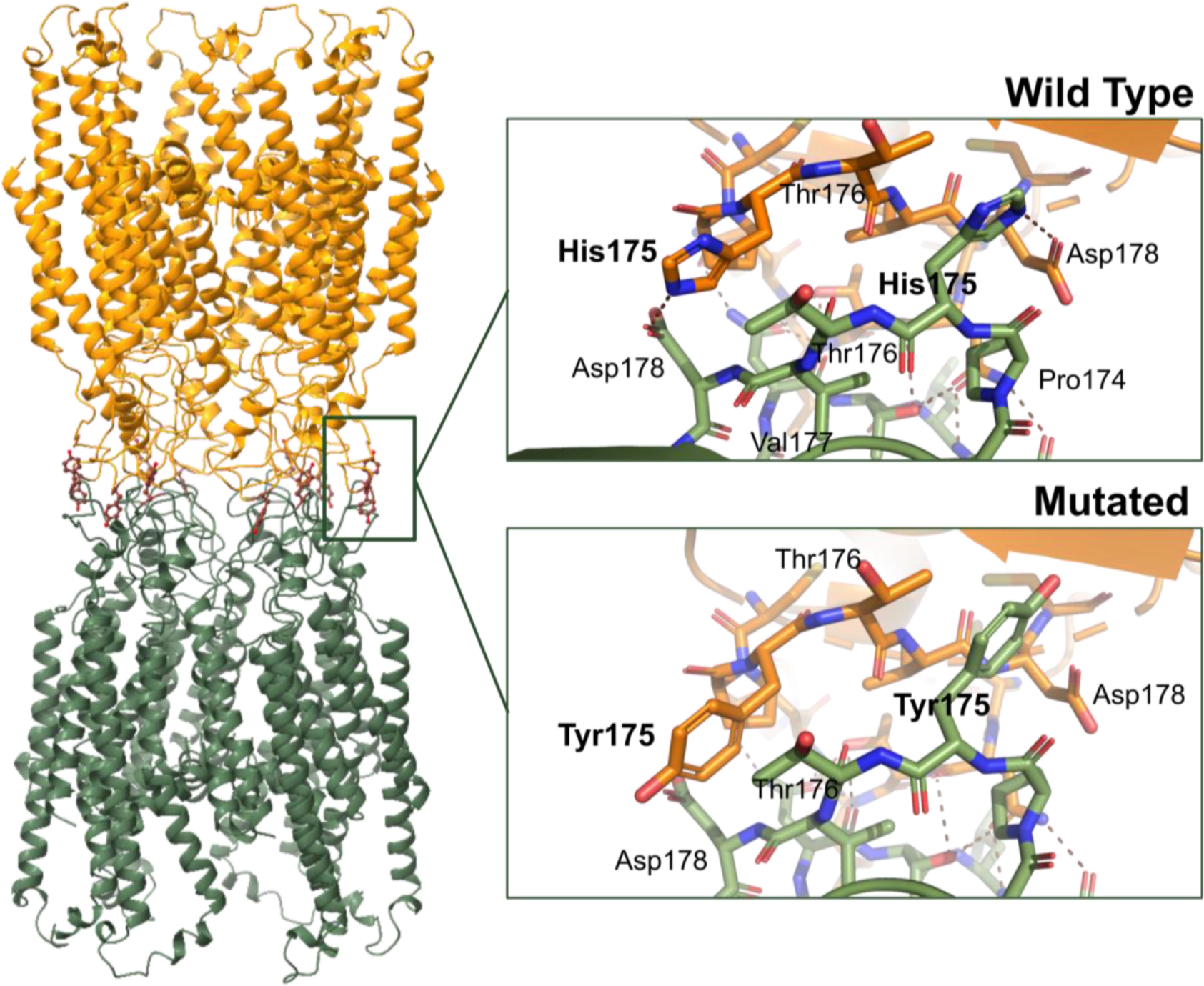
Model of the human CX31.9 gap junction formed by two homomeric connexons; and change produced by the NP_689343.3:p.(His175Tyr) variant comparing the wild-type protein with the mutated.

The NP_689343.3:p.(His175Tyr), NP_689343.3:p.(Pro248del), NP_689343.3:p.(Arg253Pro) variants were predicted in-silico as neutral (−0.5 < ΔΔG_pred_ < 0.5) according to the predicted change in global GJD3 monomer stability for the majority of methods used (Table S4). The effect on protein stability of the NP_689343.3:p.(His175Tyr) and NP_689343.3:p.(Arg253Pro) variants, found together in the same patients, have also been predicted to be neutral.

In the homomeric connexon and the homotypic gap junction models, the NP_689343.3:p.(His175Tyr) variant was predicted to have a stabilizing effect on the structure of the complex (Table S5). Nevertheless, based on the model and the interaction between the two connexons, the replacement of the histidine by tyrosine would affect the formation of the channel, since the electrostatic interaction between histidine 175 and aspartic 178 would be lost and replaced by the larger and uncharged amino acid tyrosine. Therefore, it could potentially alter the interaction between both connexons (Figure 2).

### 3.5. Cx30.2 localization in mouse inner ear

Immunofluorescence was used to examine the localization of the Gap junction delta-3 protein (Gjd3, Q91YD1) - also known as Cx30.2 - encoded by *Gjd3*, which is an orthologue of the *GJD3* human gene. Firstly, immunofluorescence labeling of Cx30.2 in P90 adult mouse inner ear showed dispersed punctiform labeling in the cochlea, localized at the spiral limbus, TM, nerve fibers, and organ of Corti; especially below the basal pole of inner hair cells (Figure 3A-B). In addition, immunofluorescence labeling was observed at the macula and crista epithelium (Figure S5). Significant differences were found between the *Gjd3* expression in mice cochleae of animals that were one and six months (*p* = 0.011); the lower value of ΔCt at 1-month-old showed a higher expression than in 6-month-old mice (FC ± SEM = 0.610 ± 0.057). Although expression was higher in 1-month-old mice than in 12-month-old (FC ± SEM = 0.747 ± 0.111), that difference was not statistically significant (*p* = 0.073, Figure 3C).

**Figure 3.**
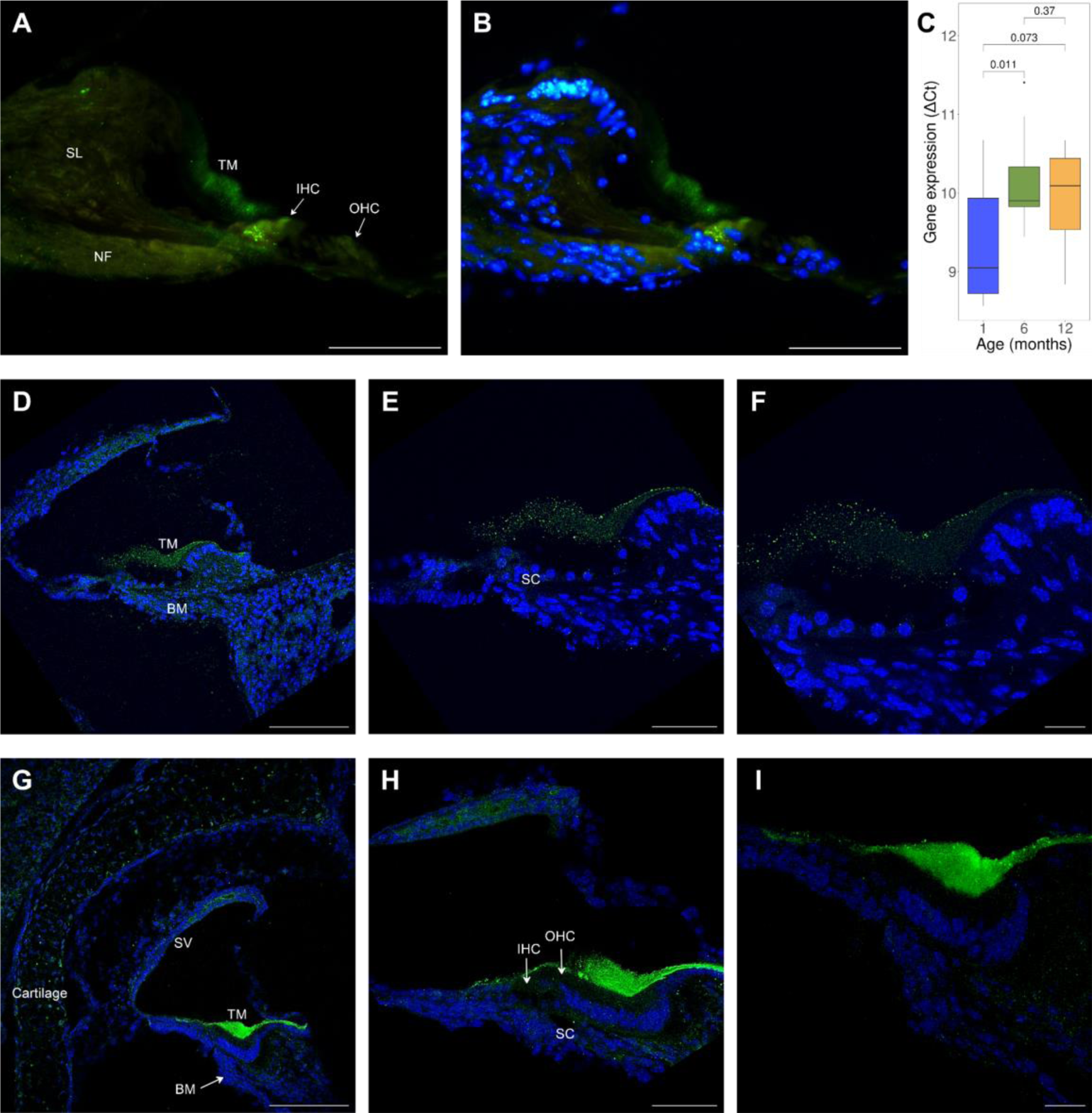
Expression of Cx30.2 at the cochlea of postnatal day 90 (P90) adult (A-B), postnatal day 30 (P30) adult (D-F), and postnatal day 3 (P3) young (G-I) mouse inner ear sections, and expression in mouse development (C). A-B: Gjd3 appears in the cochlea in a very disperse punctiform immunofluorescence labeling at spiral limbus (SL), nerve fibers region (NF), tectorial membrane (TM) and the organ of Corti; with more dense patches at TM or below the inner hair cells (IHC). C: Gene expression (ΔCt) of *Gjd3* in 1, 6 and 12-month-old mouse cochleae (in each age group, there are four pools, each consisting of three cochleae). Student’s t-test was used to calculate the *p* value in each comparison. D-F: The punctiform immunofluorescence labeling of Gjd3 in the P30 cochlea was observed especially at the TM, and also at the basilar membrane (BM) and supporting cells (SC). G-I: The immunofluorescence revealed strong expression of Gjd3 in the P3 cochlea at the TM, and also in a punctiform labeling at the cartilage, stria vascularis (SV), BM, IHC, outer hair cells (OHC), and SC. The Connexins 30.2 (Cx30.2) - encoded by the mouse *Gjd3* gene - are stained using the rabbit polyclonal anti-Connexin 30.2, LifeTechnologies, # 40-7400 (green), and the nuclei are stained using DAPI (blue). Scale bar 50 μm (A, B, E and H), 100 μm (D and G), and 20 μm (F and I).

Therefore, cochleae from younger mice were studied to analyze the localization of Cx30.2 during development. In P30 adult mouse cochleae, expression was predominantly observed in the TM but also in the supporting cells of the organ of Corti and the basilar membrane (Figure 3D-F). In the P3 young mouse cochleae, the labeling of the TM was the most intense; furthermore, Cx30.2 was expressed in the hair and supporting cells from the organ of Corti, stria vascularis, basilar membrane, and cartilage (Figure 3G-I).

### 3.6. Functional characterization of human CX31.9 in *Xenopus laevis* oocytes

To examine the functional properties of human CX31.9 hemichannels, we expressed the CX31.9 in *Xenopus laevis* oocytes, and measured currents in response to a wide range of voltage steps (+80 to - 100 mV) from a holding potential of -40 mV. Consistent with the functional expression of hemichannels, *Xenopus* oocytes injected with CX31.9 WT RNA showed significantly larger currents at +80 mV and - 100 mV compared to uninjected oocytes (*p* < 0.0001, *n* = 33 for WT, *n* = 28 for uninjected; Figure 4B). The NP_689343.3:p.(His175Tyr) variant showed WT-level current amplitudes at +80 mV (WT: 856 ± 275 µA, *n* = 33; H175Y: 845 ± 290 µA, *n* = 28; *p* = 0.88) and -100 mV (WT: -382 ± 112 µA, *n* = 33; H175Y: -366 ± 99 µA, *n* = 28; *p* = 0.79). The NP_689343.3:p.(Arg253Pro) variant, on the other hand, showed a slight increase in current amplitudes compared to WT at +80 mV (R253P: 1007 ± 406 µA, *n* = 40, *p* = 0.065) and at -100 mV (R253P: -453 ± 141 µA, *n* = 40, *p* = 0.0092).

**Figure 4.**
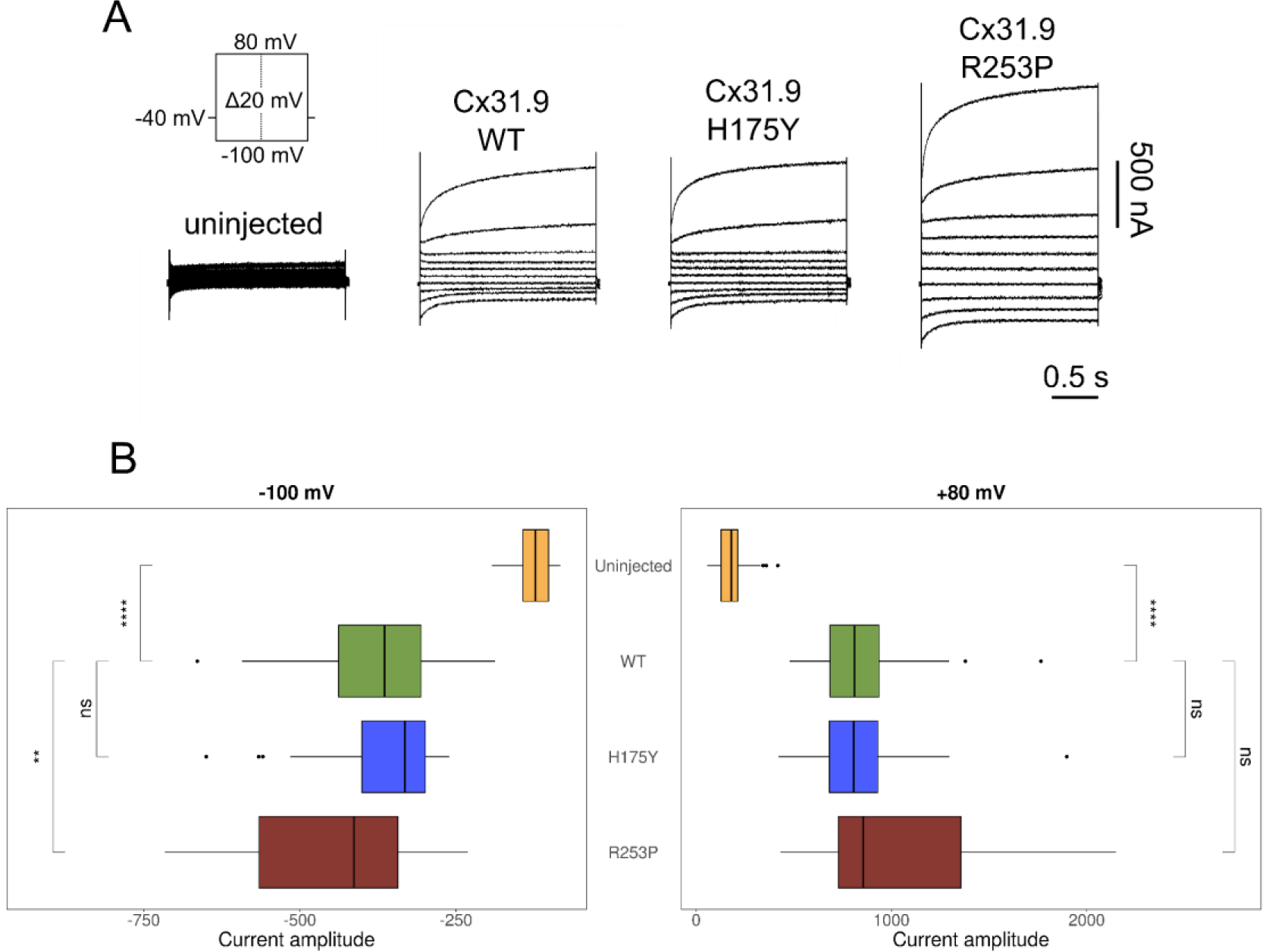
Functional expression of human CX31.9 hemichannels in *Xenopus laevis* oocytes. A: Representative current traces of *Xenopus* oocytes that are untreated (not injected with RNA) and expressing CX31.9 wild-type, NP_689343.3:p.(His175Tyr) and NP_689343.3:p.(Arg253Pro) hemichannels in response to a voltage-step protocol from +80 mV to -100 mV (holding potential = -40 mV). B: Current amplitudes elicited at +80 mV and -100 mV. Floating bars show the third quartile, median (middle line) and first quartile values. WT: wild-type; H175Y: NP_689343.3:p.(His175Tyr); R253P: NP_689343.3:p.(Arg253Pro); ns: *p* < 0.05; **: *p* ≤ 0.01; ****: *p* ≤ 0.0001; Student’s t-test.

## 4. Discussion

The main finding in this work is the burden of rare variants in the human *GJD3* connexin gene in FMD. By manual inspection and segregation analyses of these variants, our study has identified a rare TGAGT haplotype in the gene *GJD3* that segregates the complete phenotype in multiple unrelated families with MD and supports an association of *GJD3* with FMD. Furthermore, immunofluorescence experiments reveal the presence of Gjd3 protein in mice cochleae and vestibules; and, unexpectedly, Gjd3 has been localized for the first time in the TM.

Two missense, two synonymous and one downstream variants segregate with dominant pattern in three different families with some individuals affected by MD; in addition, in another two familial cases and eight sporadic individuals, having four of them first-degree relatives with incomplete phenotype. The 18 MD cases shared the same haplotype TGAGT for the variants: NC_000017.11:g.40356228C>T, NC_000017.11:g.40363058C>G, NC_000017.11:g.40363293G>A, NC_000017.11:g.40363294C>G and NC_000017.11:g.40363579G>T; whose frequency in the Iberian population in Spain is 0.0093. The low frequency of the haplotype and the segregation in non-related families leads to the association with the disease. The most interesting variant is NC_000017.11:g.40363293G>A, NP_689343.3:p.(His175Tyr) at the protein level, which produces an amino acid change from a positively charged histidine to a bulky and hydrophobic tyrosine in the extracellular extreme of the connexon. This replacement would produce the loss of the electrostatic interactions that occur between histidine 175 and aspartic 178 in each of the six connexins conforming the homomeric connexon, altering the correct arrangement between two connexons to form the channel. Although the electrophysiological characterization of WT and the mutated NP_689343.3:p.(His175Tyr) CX31.9 hemichannels in *Xenopus laevis* oocytes demonstrated no differences in the current amplitudes (Figure 4B); the amino acid change could modify the interaction between both connexons, leading to decreased formation of homotypic gap junction channels. Likewise, Schadzek et al. ^59^ demonstrated that a missense variant in an extracellular loop (as in our case) of CX46 - encoded by *GJA3* - is related to an autosomal dominant zonular pulverulent cataract. In this case, they demonstrated that the mutated connexin affected the co-expressed wild type (wt) connexin to achieve a dominant inheritance. The heterodimer mutated-wt made less gap junction plaques than the homodimer wt-wt, and the homodimer mutated-mutated formed almost none.

In addition, it has been demonstrated that Gjd3 can form heterotypic channels with other three connexins in mice heart cells ^60,61^. We suspect that, in the same way, Gjd3 could form heteromeric connexons and heterotypic channels in the cochlea with the other connexins expressed, as it has been demonstrated with Cx26, Cx30 and Cx31 - also named Gjb2, Gjb6 and Gjb3, respectively. Heteromeric Gjb2/Gjb6 channels have been found connecting cochlear supporting cells, and the Gjb2 and Gjb3 also form heteromeric Gjb2/Gjb3 connexons and homomeric/heterotypic Gjb2/Gjb3 gap junctions ^62–64^. The correct arrangement of a heterotypic connexon also would be affected by the NC_000017.11:g.40363293G>A variant. Nevertheless, it cannot be asserted that the expression of human GJD3 is identical to that of mouse Gjd3 in the cochlea, as observed in the heart ^65^.

It is currently not possible to model the mutant hemichannel NP_689343.3:p.(Arg253Pro) as the residue is located in the highly flexible cytoplasmic region, which also remains unresolved in other connexin structural studies ^66^. Therefore, we used an electrophysiological assay to assess potential functional impact of this variant. Electrophysiological characterization of the mutated NP_689343.3:p.(Arg253Pro) CX31.9 hemichannel in *Xenopus laevis* oocytes showed that at hyperpolarized potentials, the current amplitudes were significantly larger than the WT hemicannels (Figure 4B). As the increase in current amplitudes was small and the functional significance of the cytoplasmic region of connexins is currently poorly understood, we refrain from drawing any conclusion about this finding. Future studies investigating the effect of this variant in gap junction channels will help clarify the pathogenicity of this variant.

Hearing relies on the displacements of the stereocilia of hair cells provoked by sound. The membrane depolarization entails fluxes of Ca^2+^ and K^+^ into the cell, leading to the excitation of the auditory nerve ^67,68^. As evidenced in prior research, inner ear connexins are essential in the Ca^2+^ signaling ^69^. Besides, there are Ca^2+^-rich filamentous structures in the TM that are involved in the connection of the TM and the hair cell stereocilia, which assure the mechanical stimulation and the obtention of Ca^2+^ by the hair cells ^68^. Our IF data confirm the expression of Gjd3 in the mouse TM and we speculate that GJD3 could be related to the function of those filamentous ducts. Moreover, regarding the cycling transport of K^+^, the K^+^ flows through the hair cells from the endolymph to the perilymph ^70^. The TM is sealed, therefore to arrive at the hair cells from the endolymph, the K^+^ must cross the TM ^71^. We hypothesize that GJD3 may contribute to maintaining the local ionic microenvironment driving K^+^ fluxes to the tip of stereocilia. When the K^+^ arrives to the hair cells, it reaches to the perilymph through scala tympani, then to the spiral ligament, and arrives to the stria vascularis, where it is returned to the endolymph. It has been studied that the GJB2, GJB3 and GJB6 connexins are crucial in this transport ^4,63,70^. Because of that, we propose that GJD3, which we found expressed some of these cells and structures, should be involved in the K^+^ cycle.

Furthermore, the connexins Gjb2, Gjb6 and Gja1 have been found in mammalian vestibular system ^72^. Our IF data in the vestibular organs confirm a labeling of Gjd3 below the macular epithelium; however, further studies with higher resolution are needed to confirm these observations.

By exome sequencing and familial analysis different genes have been found associated with MD. Particularly, an enrichment of rare missense variants in 15 unrelated MD families, with 6 of them showing compound heterozygous recessive inheritance in *OTOG*, and rare missense variants and two short deletions were identified in four different MD families in *TECTA* genes, respectively; both genes encode non-collagenous proteins of the TM ^13,15^. Moreover, the other nine unrelated families presented rare variants in the *MYO7A* gene, expressed in the stereocilia of the hair cells in the sensory epithelia ^14^. Taken together, these studies and the findings in *GJD3,* suggest that the proteins involved in the architecture of the stereocilia links and the attachment of the stereocilia tips to the TM could be involved in the pathophysiology of MD.

In the present work, the type of inheritance observed in the TGAGT rare haplotype in *GJD3* in three MD families was autosomal dominant. Three different inheritance modes have been reported in FMD: autosomal dominant, autosomal recessive and digenic inheritance. These outcomes describe a complex inheritance, that coupled with specific environmental factors, could lead to variation of phenotype, including the HL profile, and age of onset, even in the same family ^73^. Moreover, epigenetic modifications could probably shape clinical manifestation. By whole-genome bisulfite sequencing (WGBS), CpGs in *ADGRV1* (MIM: 602851), *CDH23* (MIM: 605516) and *PCDH15* (MIM: 605514) were determined as differentially methylated when comparing MD against healthy controls. Those genes encode for stereocilia link proteins, which are involved in attaching the hair cells to the TM ^11^.

Furthermore, in the 3 non-familial cases with the CGGCG haplotype (displaying the reference alleles except for the NC_000017.11:g.40363058C>G missense variant), the variant alone would not explain the phenotype. However, it is very interesting for future work opening the possibility to study the complete genome to identify regulatory variants in *GJD3,* as in the promoters or in the 5’ or 3’ untranslated regions (UTRs); and/or study in combination the genetics with epigenetics in sporadic cases.

## Conclusions

1. A rare haplotype in the gene *GJD3* segregates in unrelated families with Meniere Disease.
2. Cx30.2 is localized in mouse cochlea, including the tectorial membrane, and vestibule.
3. The variants found may involve the interactions between two connexons leading to dysfunction in the channels.
4. In line with previous findings, our results support that the proteins of the tectorial membrane and the stereocilia link could be involved in the molecular pathophysiology of familial MD.

## Limitations

1. The lack of clinical record and DNA samples of some participants, made it difficult to segregate the haplotype in some individuals.
2. This study was limited to coding sequences, and whole genome sequencing data will be needed to study non-coding regions and its relation with the disease.
3. Additional functional studies will be necessary to understand the function and the consequences of the missense variants in the human CX31.9 protein in the inner ear.

## Acknowledgments

This work is part of Alba Escalera-Balsera’s doctoral thesis. Alba Escalera-Balsera is enrolled in the Biomedicine Ph.D. program at the University of Granada, Spain. We would also like to recognize Dr Lidia Frejo for suggestions with immunofluorescence analysis.

## Author contributions

Conceptualization, A.E.-B. and J.A.L.-E.; Methodology, A.E.-B., P.R.-B., A.M.P.-P., C.H.C., L.R.- dlR., J.C. and E.D.; Software, A.E.-B., A.M.P.-P. and A.G.-M.; Formal Analysis, A.E.-B.; Investigation, A.E.-B., S.M.-C., C.H.C. and A.G.-M.; Resources, J.C.A.-D., A.S.-V., I.V.-N., A.J.S. and J.A.L.-E.; Data Curation, A.E.-B.; Writing – Original Draft Preparation, A.E.-B. and J.A.L.-E.; Writing – Review & Editing, P.R.-B., A.M.P.-P., S.M.-C., C.H.C., L.R.-dlR., J.C., E.D., J.C.A.-D., A.S.-V., I.V.-N., A.J.S., A.G.-M. and J.A.L.-E.; Visualization, A.E.-B., P.R.-B. and A.M.P.-P.; Supervision, I.V.-N., A.J.S., A.G.-M. and J.A.L.-E.; Project Administration, J.A.L.-E.; Funding Acquisition, J.A.L.-E..

## Institutional review board

The study was conducted according to the guidelines of the Declaration of Helsinki and approved by the Granada Ethical Review Board for Clinical Research under the protocol number 1805-N-20; the Governmental Ethics Commission for Animal Welfare in Berlin under the approval number T 0235/18; and by the Dirección General de Agricultura, Ganadería y Alimentación in Comunidad de Madrid under the approval number PROEX 325.4/21.

## Informed consent

Written informed consent has been obtained from the patient(s) to publish this paper.

## Data availability

Anonymized genetic raw dataset and family pedigrees used in this study are available from the corresponding author upon reasonable request.

