## Supplementary material for "A rare haplotype of the *GJD3* gene segregating in familial Meniere Disease interferes with connexin assembly"

#### Supplementary figures

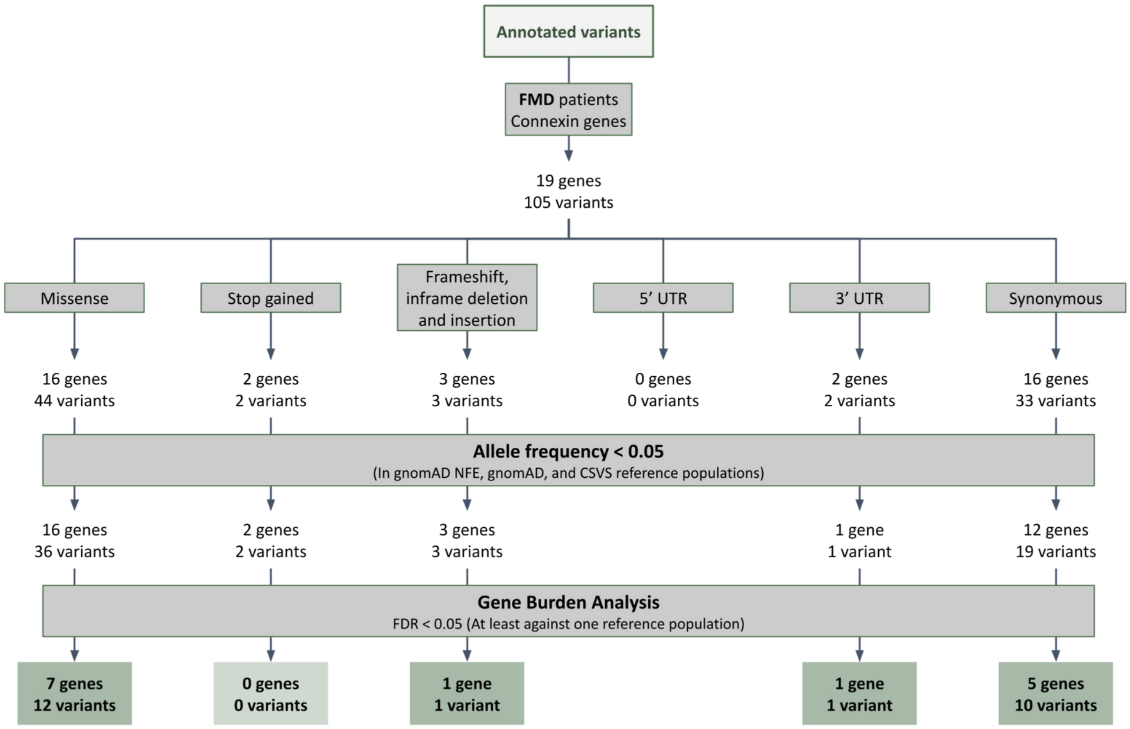


**Figure S1.** Flow chart summarizing the prioritization strategy and the result of the Gene Burden Analysis for Familial Meniere Disease (FMD) patients. FMD: Familial Meniere Disease; UTR: Untranslated regions; gnomAD NFE: Non-Finnish European for gnomAD; gnomAD: Global population for gnomAD; CSVS: Collaborative Spanish Variant Server, Spanish population; FDR: p-value corrected by False Discovery Ratio.

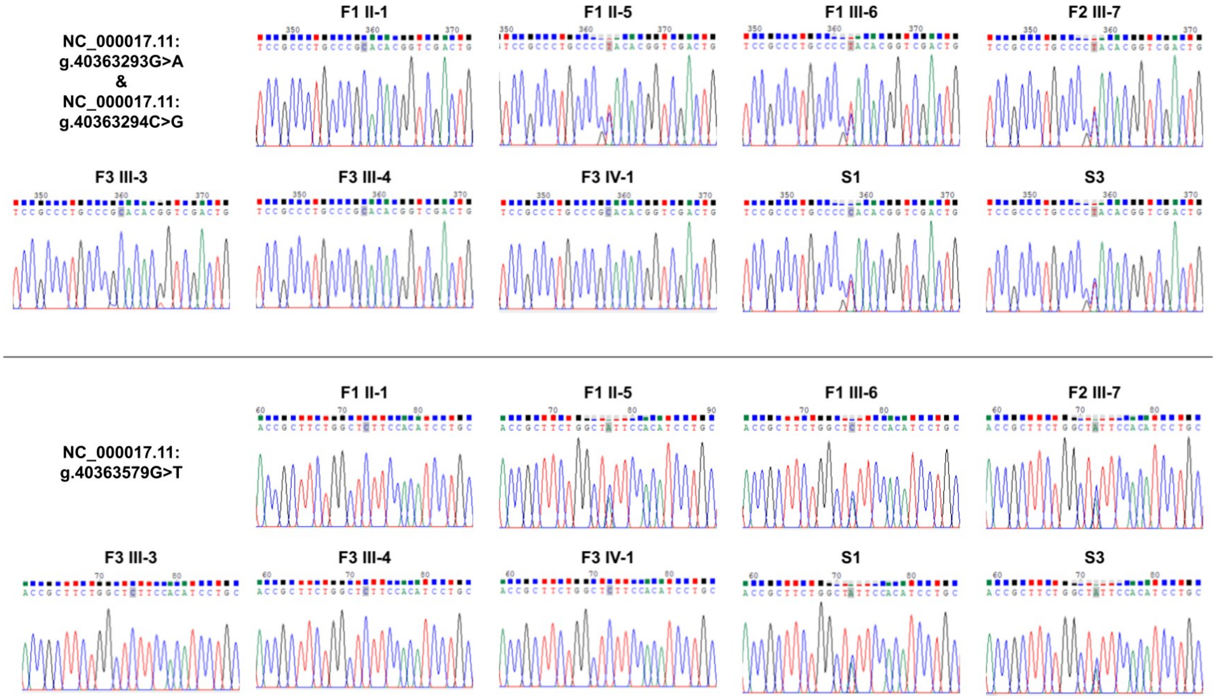


**Figure S2.** Results of Sanger Sequencing for the NC_000017.11:g.40363293G>A, NC_000017.11:g.40363294C>G and NC_000017.11:g.40363579G>T variants in *GJD3.* For the samples F1 II-1, F1 II-5, F1 III-6, F2 III-7, F3 III-3, F3 III-4, F3 IV-1, S1 and S3.

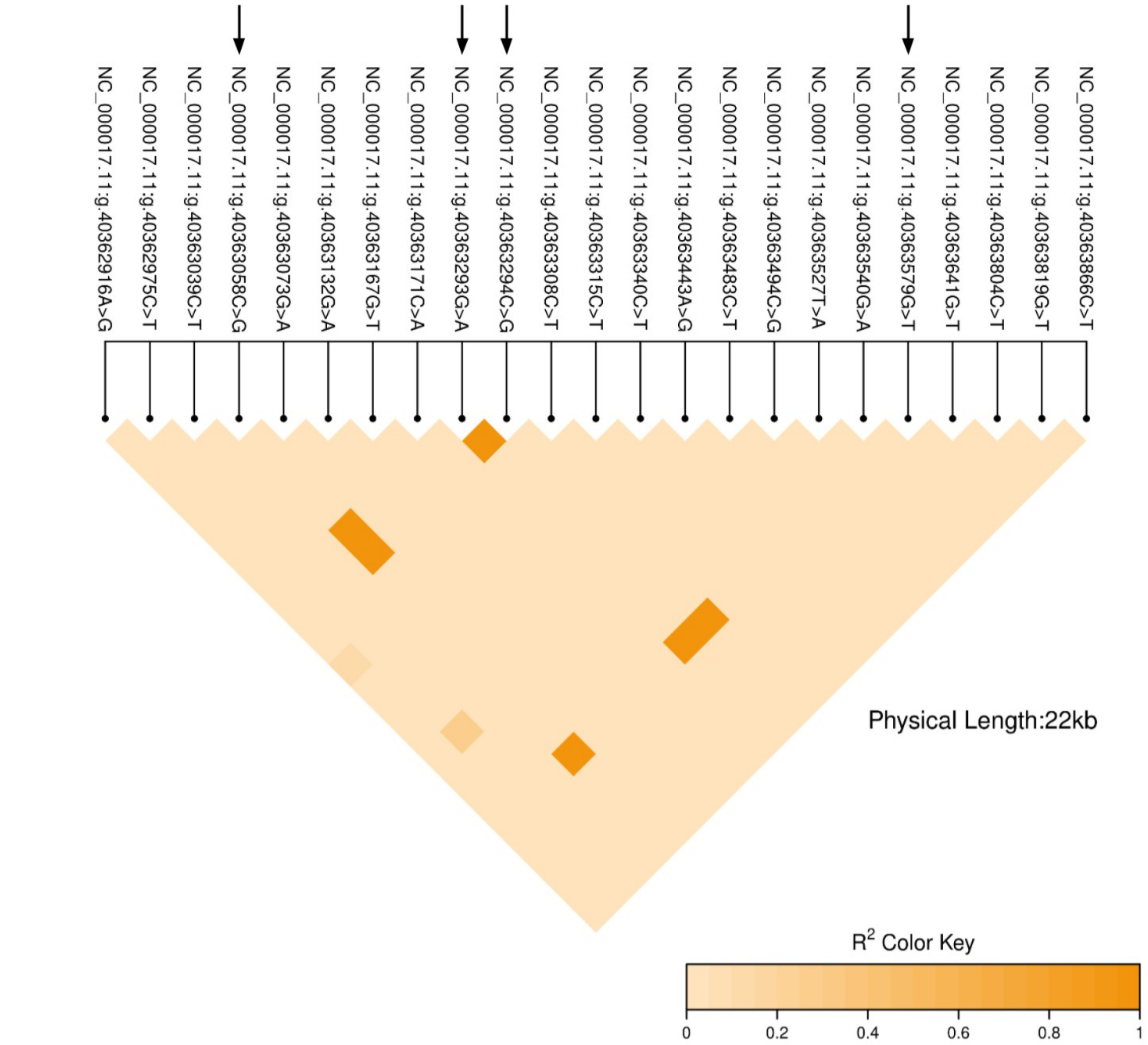


**Figure S3.** Heatmap representing a pairwise linkage disequilibrium of variants in the *GJD3* gene. The variants tagged with an arrow are in the same block, and the variants are shared by the individuals studied.

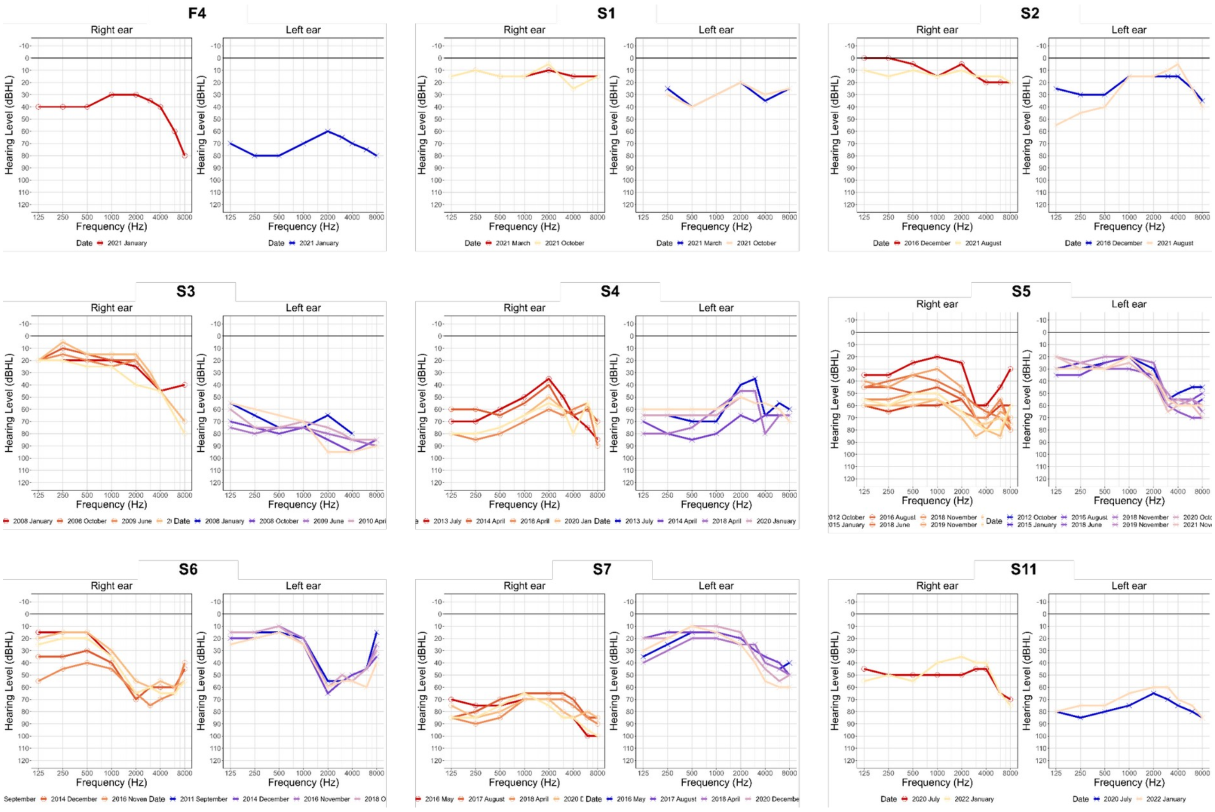


**Figure S4.** Air conduction audiogram of F4, S1, S2, S3, S4, S5, S6, S7 and S11 individuals. dB: decibels, kHz: kilohertz.

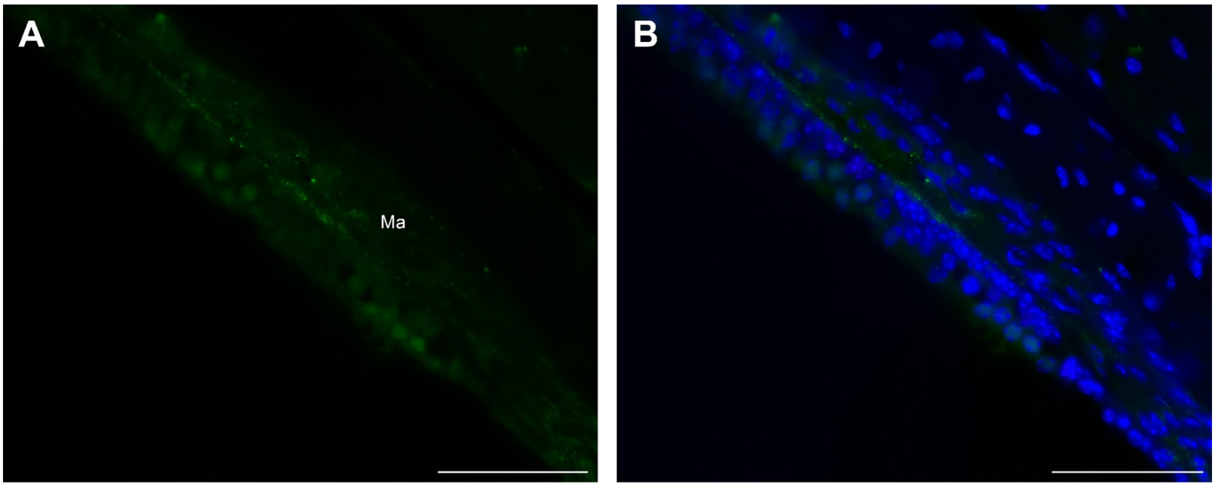


**Figure S5.** Presence of Gjd3 at the cochlea of postnatal day 90 (P90) adult mouse inner ear sections. Gjd3 immunolabeling is localized at the macula (Ma) and crista of the vestibulum (data not shown). The Connexins 30.2 (Cx30.2) - encoded by the mouse *Gjd3* gene - are stained using the rabbit polyclonal anti-Connexin 30.2, LifeTechnologies, # 40-7400 (green), and the nuclei are stained using DAPI (blue). Scale bar 50 µm.

### Supplementary tables

**Table S1.** Clinical and demographic variables assessed in the familial Meniere Disease (FMD) individuals.

| | FMD (n = 70) |
|---|---|
| **Sex (% woman)** | 61.54 |
| **Age (mean±SEM)** | 59.6±1.45 |
| **Age of onset (mean±SEM)** | 36.53±1.62 |
| **Hearing loss laterality (% bilateral)** | 48.39 |
| **Headache (% yes)** | 35.19 |
| **Type headache (% migraine)** | 52.38 |
| **History of autoimmune disease (% yes)** | 27.50 |
| **PTA (mean±SEM)** | 47.08±2.1 |
| **4,000Hz (mean±SEM)** | 53.13±2.57 |
| **8,000Hz (mean±SEM)** | 61.07±2.49 |

SEM: Standard error of the mean; PTA: Pure tone audiogram, calculated as the mean of the 500Hz, 1,000Hz and 2,000Hz frequencies; Hz: Hertz.

**Table S2.** Allelic frequencies (AF) of the studied haplotype in our Meniere Disease (MD) cohort and in the 1,000 Genomes Project cohorts.

| Haplotype name | NC_000017.11:g. | | | | | AF MD cohort | | AF 1,000 Genomes Project | | |
|---|---|---|---|---|---|---|---|---|---|---|
| | **40356228** | **40363058** | **40363293** | **40363294** | **40363579** | **FMD in GBA** | **SMD** | **AF ALL** | **AF EUR** | **AF IBS** |
| Reference | C | C | G | C | G | 9.29E-01 | 9.65E-01 | 9.94E-01 | 9.83E-01 | 9.86E-01 |
| TGAGT | T | G | A | G | T | 7.14E-02 | 2.56E-02 | 5.80E-03 | 1.59E-02 | 9.30E-03 |
| CGGCG | C | G | G | C | G | 0.00E+00 | 9.58E-03 | 2.00E-04 | 1.00E-03 | 4.70E-03 |

AF: Allelic frequency; FMD: Familial Meniere Disease; GBA: gene burden analysis; SMD: Sporadic Meniere Disease; ALL: global population; EUR: European population; IBS: Iberian population from Spain.

**Table S3.** Model validation of predicted 3D protein structural models.

| Protein | Modeled length | Modeling method | Evaluation | | | | |
|---|---|---|---|---|---|---|---|
| | | | Molprobity Score | Verify3D | ERRAT | ProSA-Web | QMEANDisCo |
| GJD3 monomer | 294 | AlphaFold2 | 1.8 | 40.14 | 87.6106 | -4.66 | 0.56 ± 0.05 |
| GJD3 hemicha-nnel | 6 chains of 209 residues | AlphaFold2 and HDOCK | 0.84 | 26.40 | 92.0067 | -5.57 | 0.63 ± 0.05 |
| GJD3 channel | 12 chains of 209 residues | AlphaFold2 and HDOCK | 0.84 | 26.28 | 90.8372 | -5.57 | 0.63 ± 0.05 |

Molprobity Score, Verify3D, ERRAT, ProSA-web and QMEANDisCo were used to evaluate the quality of the predicted structural models. Molprobity Score is a weighted logarithmic combination of geometric scores such as clashscore, percentage of unfavored Ramachandran and bad sidechain rotamers. Lower values indicate better quality.

Verify3D assesses the compatibility of an atomic model with its amino acid sequence. ERRAT analyzes interactions between atoms and provides an overall quality factor. Models with values >50 revealed that the backbone conformation and nonbonded interactions of all models were within the scope of a high-quality model. ProSA-web uses a z-score to measure the energy separation between the native fold and the average of an ensemble of the misfolds in standard deviation units of the protein database. QMEANDisCo evaluates agreement of pairwise distances between residues and distance constraints from homologous structures. Higher scores indicate higher quality models.

**Table S4.** Protein stability change prediction caused by the found variants in the GJD3 model.

| Predictor | Protein variant | | | NP_689343.3: p.(His175Tyr) + NP_689343.3 :p.(Arg253Pro) |
|---|---|---|---|---|
| | **NP_689343.3: p.(His175Tyr)** | **NP_689343.3: p.(Pro248del)** | **NP_689343.3: p.(Arg253Pro)** | |
| ENCoM (kcal/mol) | Neutral (0.130) | Neutral (0.056) | Destabilizing (-0.721) | - |
| SDM (kcal/mol) | Neutral (-0.110) | Stabilizing (0.630) | Destabilizing (-1.310) | - |
| mCSM (stability) (kcal/mol) | Stabilizing (1.21) | Neutral (-0.403) | Neutral (0.237) | - |
| mCSM-membrane (kcal/mol) | Destabilizing (-0.519) | Neutral (-0.258) | Neutral (-0.255) | - |
| DynaMut2 (kcal/mol) | Stabilizing (1.410) | Neutral (-0.14) | Neutral (-0.010) | Neutral (0.31) |
| PremPS (kcal/mol) | Neutral (0) | Neutral (0.11) | Neutral (0.050) | - |
| I-Mutant | Neutral (-0.1) | Neutral (-0.3) | Neutral (0.1) | Neutral (0.1) |

For PremPS, $\Delta\Delta G_{pred} < 0.0$ indicates a stabilizing mutation although for ENCoM, DynaMut2, I-Mutant, mCSM, mCSM-membrane and, SDM $\Delta\Delta G_{pred} > 0.0$ indicates a stabilizing mutation. Mutations are classified as neutral mutations when $-0.5 < \Delta\Delta G_{pred} < 0.5$.

**Table S5.** Protein stability change prediction caused by NP_689343.3:p.(His175Tyr) variant in the GJD3 hemichannel and channel models.

| Predictor | GJD3 Hexamer | GJD3 Dodecamer |
|---|---|---|
| DynaMut2 (kcal/mol) | Stabilizing (1.6) | Stabilizing (1.6) |
| mmCSM-PPI (kcal/mol) | Stabilizing (0.66) | Stabilizing (0.94) |

Global protein stability change prediction (kcal/mol) using two different ΔΔGpred prediction methods. For DynaMut2 and, mmCSM-PPI ΔΔGpred > 0.0 indicates a stabilizing mutation. Mutations are classified as neutral mutations when -0.5 < ΔΔGpred < 0.5.
